# Predicting Acute Cerebrovascular Events in Stroke Alerts Using Large-Language Models and Structured Data

**DOI:** 10.1101/2025.09.28.25336852

**Authors:** Asala N Erekat, Margaret H Downes, Laura K Stein, Bradley N Delman, Adam M Karp, Ankita Tripathi, Girish N Nadkarni, Mark J Kupersmith, Benjamin R Kummer

## Abstract

**Background:** Acute stroke alerts are often activated for non-cerebrovascular conditions, leading to false positives that strain clinical resources and promote diagnostic uncertainty. We sought to develop machine learning (ML) models integrating large-language models (LLMs), structured electronic health record data, and clinical time series data to predict the presence of acute cerebrovascular disease (ACD) at stroke alert activation.

**Methods:** We derived a series of ML models using retrospective data from stroke alerts activated at Mount Sinai Health System between 2011 and 2021. We extracted structured data (demographics, medical comorbidities, medications, and engineered time-series features from vital signs and lab results) as well as unstructured clinical notes available prior to the time of stroke alert. We processed clinical notes using three embedding approaches: word embeddngs, biomedical embeddings (BioWordVec), and LLMs. Using a radiographic gold standard for acute intracranial vascular event, we used an auto-ML approach to train one model based on unstructured data and five models based on different combinations of structured data. We evaluated models individually using the area under the receiver operating characteristic curve (AUROC), mean positive predictive value (PPV), sensitivity, and F1-score. We then combined the 6 model logits into a multimodal ensemble by weighting their logits based on F1-score, determining ensemble performance using the same metrics.

**Results:** We identified 16,512 stroke alerts corresponding to 14,233 unique patients over the study period, of which 9,013 (54.6%) were due to ACD. The multi-modal model (AUROC 0.72, PPV 0.68, sensitivity 0.76, F1 0.72) outperformed all individual models by AUROC. One structured model based on demographics, comorbidities, and medications demonstrated the highest sensitivity (0.95).

**Conclusions:** We developed a multi-modal ML model to predict ACD at stroke alert activation. This approach has promise to optimize stroke triage and reduce false-positive activations.

## Introduction

Acute cerebrovascular events, including acute ischemic stroke (AIS), intracerebral hemorrhage (ICH), subdural hematoma (SDH), and subarachnoid hemorrhage (SAH), are leading causes of morbidity and mortality worldwide.^1–4^ Stroke alert systems are designed to rapidly identify cerebrovascular events and facilitate timely treatment interventions such as thrombolysis or mechanical thrombectomy.^5^ However, these systems face significant challenges. Up to 67% of alerts are “false positive” alerts, which are often triggered by ‘stroke mimic’ conditions including sepsis, metabolic abnormalities, or respiratory failure.^6–8^ Additionally, up to one-third of patients hospitalized with stroke-like symptoms do not actually ultimately receive a cerebrovascular diagnosis.^9^

High false-positive alerts can lead to inefficient resource allocation, increases in hospital length-of-stay, and delays to appropriate care for both stroke and non-stroke patients. Developing a method of distinguishing acute cerebrovascular disease (ACD) from non-stroke entities could address this problem from two perspectives: (1) to accelerate and increase diagnostic precision in patients for whom a stroke alert has been activated, or (2) to potentially prevent the activation of stroke alerts before they are triggered.

The increasing clinical and economic burden of stroke has driven interest in leveraging machine learning (ML) and artificial intelligence (AI) to improve the specificity of stroke alert systems.^10^ In recent years, large-language models (LLMs) have emerged as a powerful tool for extracting clinically relevant insights from unstructured free-text data, which often contains nuanced information about patient presentations that structured data alone may not capture.^11–14^ LLMs are therefore a promising method for developing predictive models to enhance stroke alert accuracy.

Prior efforts at accomplishing this, or similar, tasks have notable limitations. While one prior study developed a predictive model based on audio and video data of patients with suspected stroke, these data types are not commonly obtained during stroke alert evaluations.^15^ Other approaches at accelerating early detection of acute stroke with ML were limited by small populations or cohorts with disproportionately high prevalence of true cerebrovascular disease,^16–18^ by a focus on only specific stroke types,^19^ or by reliance on narrow sets of structured features as inputs.^17,20^ Most studies were also restricted to emergency department (ED) settings only,^15–17,19,20^ rather than both ED and inpatient hospitalizations.

In addition, while recent approaches have made use of natural-language processing (NLP) techniques to include unstructured clinical text data in predictive ML models,^19^ these approaches have not leveraged LLM methods. Finally, recent efforts have not developed models specifically for patients with activated stroke alerts. Instead, they have relied on hospital presentation or ED arrival as the prediction timepoint, rather than the moment when acute cerebrovascular disease (ACD) was first suspected.^15–20^ This is a limitation for developing models to improve the yield for such events without sacrificing sensitivity.

In light of these challenges, we sought to develop multiple predictive ML models using a large dataset of stroke alerts from an urban academic hospital network (Figure 1). We employed two primary data modalities: (1) structured electronic health record (EHR) data available at the time of stroke code activation for which distinct models were trained to capture various components; and (2) unstructured clinical notes, where models based on LLM embeddings were used to capture the subtle and context-rich documentation related to stroke presentations. We hypothesized that a multi-modal model incorporating both data modalities would demonstrate superior performance in discriminating true acute cerebrovascular events from “false-positive” alerts than either text-based or structured-data only models alone.

**Figure 1.**
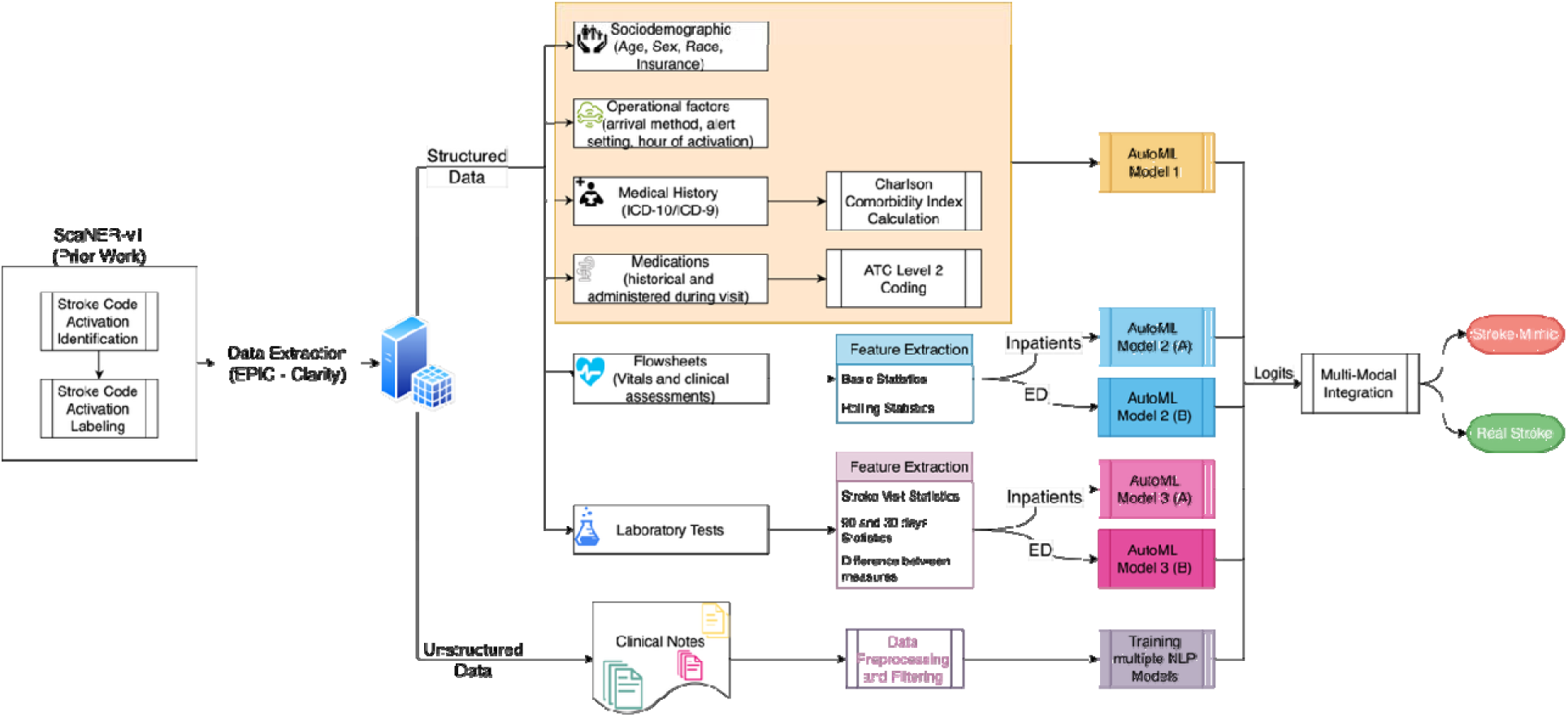
Overview of data processing and multi-modal modeling pipeline. Structured and unstructured EHR data were extracted for hospital encounters in which stroke alerts were activated. Structured inputs included demographics, comorbidities, medications, labs, and vitals; unstructured inputs were clinical notes. Separate models were trained for different data modalities, and their outputs were integrated into a final ensemble to classify stroke vs. stroke mimic at the time of alert activation.

## Methods

### Patient Population

We derived a series of ML models using retrospective-collected data from patients for whom stroke alerts were activated at the Mount Sinai Health System (MSHS), a 7-site academic medical center in New York City. Using our institutional clinical data warehouse, we identified all stroke alerts that were activated at all MSHS campuses between Jan 1, 2011, and June 2021, with the start of the follow-up period coinciding with the go-live of the current EHR (Epic, Epic Systems Corporation, Verona WI, USA) at MSHS. Each stroke alert was labeled with a binary outcome (presence or absence of ACD) based on the output of ScanNER, a NLP framework that applied rule-based named-entity recognition (NER) to stroke alert-associated neuroradiology reports and combined this with validated stroke diagnosis code algorithms. Details of this framework, including sample size calculations, are included in forthcoming research work from our laboratory (Asala N. Erekat, PhD, unpublished data, 2025).

This study was approved by the Institutional Review Board of the Icahn School of Medicine at Mount Sinai, with a waiver of informed consent due to the retrospective nature of the study and minimal risk to participants. All data were de-identified prior to analysis. The first author had full access to all the data in the study and takes responsibility for the integrity of the data and the accuracy of the data analysis.

### Data Availability

The data used in this study are not publicly available due to institutional restrictions and patient privacy considerations. De-identified data may be made available upon reasonable request to the corresponding author, subject to institutional approval. The analytic code is publicly available at https://github.com/MSHS-CNIC/ScaNER-02.

### Data Extraction

For each hospital encounter in which a stroke alert was activated, we used our institutional data warehouse to extract all unstructured clinical notes (irrespective of author or note type) documented during the hospital encounter up until the stroke alert activation time. Any notes signed after the stroke alert activation timestamp were excluded. In addition, we collected note type and author type as discrete metadata variables.

From the same source, we extracted structured EHR variables including sociodemographic factors (age, sex, race, ethnicity, insurance payor) and operational details specific to the stroke alert activation visit (means of hospital arrival, setting of stroke alert activation, hour of stroke alert). We retrieved the complete medical history available in the patient’s EHR, including all historical diagnoses coded using the *International Classification of Diseases, 9th or 10th Edition, Clinical Modification* (ICD-9-CM/10-CM). We also extracted medications administered during the hospital visit prior to the stroke code activation, along with medications documented in the patient’s historical medication list. Historical medications included active prescriptions as well as those discontinued within three months preceding the stroke activation event.

In addition, we extracted vital sign data (documented as flowsheets) from the time of hospital arrival continuously until the stroke alert activation timestamp. These included standard physiological metrics such as blood pressure, heart rate, temperature, oxygen saturation, Emergency Severity Index (ESI; a widely-accepted measure of clinical acuity), and pain scale assessments. Finally, we extracted comprehensive laboratory test results from the 90-day window preceding the stroke alert activation event, ensuring a complete contextual understanding of the patient’s recent clinical history and potential comorbidities. We focused on the particular variables that were available at the time of stroke alert activation, making them suitable for creating a model intended to support real-time detection. Figure 2 shows the different time points for each one of the extracted data sets relative to the time of stroke code activation.

**Figure 2.**
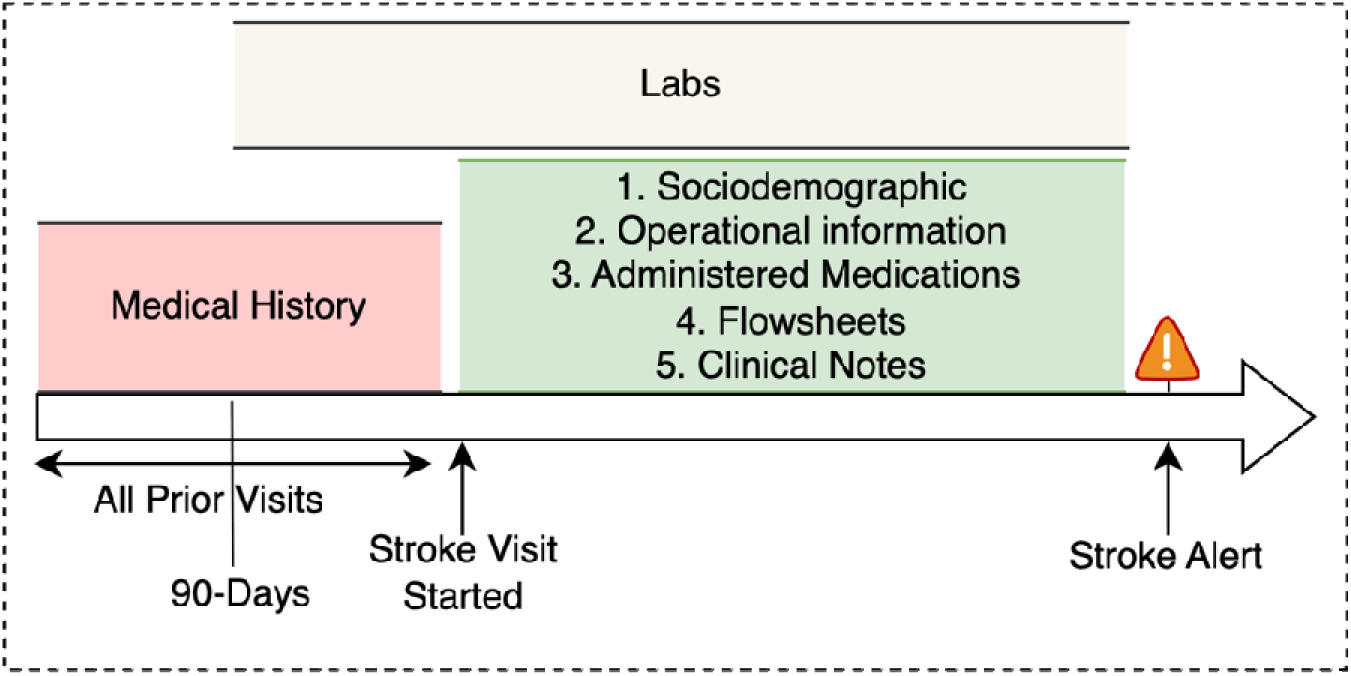
Timeline of data extraction relative to stroke alert activation. This figure illustrates the distinct time windows used to extract different data types prior to stroke alert activation. Medical history was drawn from all prior encounters while laboratory data was pulled from the preceding 90 days. Other features, including sociodemographics, operational factors, medications, flowsheets, and clinical notes, were extracted from the current stroke encounter up to the moment of alert activation.

### Data Split

To avoid data leakage, we applied a patient-level train-test split, assigning 80% of unique patients to a training set and 20% to a held-out test set. All encounters from the same patient were kept within the same group. This partition was applied consistently across all structured and unstructured modeling pipelines, ensuring that downstream comparisons and ensemble integration remained valid.

### Model Development – Unstructured Text

To transform unstructured note text into “structured” format for ML model ingestion, we used word embeddings, a widely-used technique in biomedical NLP. Word embeddings translate words as numerical arrays (or vectors) that reflect meaning, placing similar-meaning words closer together in a multi-dimensional space. Word embeddings are best conceptualized via an analogy to a postal system, wherein each word is given an “address” and words with related meanings or frequent associations are assigned “addresses” that are mathematically “close” in the semantic space. We tested three models to represent clinical note information and predict our outcome of choice: Clinical-LongFormer (CLF),^21^ MxBai,^22^ and BioWordVec.^23^

CLF is a domain-specific large language model pre-trained on medical text and fine-tuned with a built-in classification head. As CLF’s architecture is fixed, we used it as a standalone classifier to predict whether a patient’s notes indicated ACD or not, without modifying or extracting intermediary embeddings. In contrast, MxBAI is a general-purpose LLM that generates fixed-length vector representations for each clinical note. To obtain a single patient-level embedding, we pooled all MxBAI note embeddings by three strategies: mean pooling, max pooling, and a combination of both mean and max. The resulting vectors were used as input to a feedforward neural network (FNN) with two fully connected layers to minimize overfitting. We conducted one experiment for each pooling method (total three experiments), selecting the pooling strategy that generated the highest area under the receiver operating characteristic curve (AUROC) with respect to the binary ACD outcome.

In addition to CLF and MxBAI, we used BioWordVec, a static word embedding model trained on biomedical text. Unlike transformer-based models, BioWordVec assigns a consistent embedding to each word, regardless of context. For each note, we applied the same three pooling strategies as for MxBAI to summarize the word-level embeddings into a note-level vector. These were then classified using logistic regression.

For all LLM-based models, we accounted for the heterogeneity of clinical notes by training each text model (CLF, MxBAI, and BioWordVec) using 12 different combinations of author types and timings (Supplementary Table 1). Clinical notes from both ED and inpatient encounters were pooled for these analyses (Supplementary Table 2). All LLM-based models were trained on the same patient-level split described above, with performance evaluated on the held-out test set and hyperparameters optimized using a separate validation subset.

### Model Development - Structured Data

We extracted structured data available prior to stroke alert activation from our EHR, and modeled these data independently from textual features to assess their standalone predictive value. Structured data included sociodemographic characteristics, clinical information (medical comorbidities, medications, laboratory results), operational data, and physiologic time-series data. We modeled structured data using three separate modeling approaches based on data type and clinical context (ED vs. inpatient setting). Sociodemographic, medical history, and medication data were pooled across both ED and inpatient settings, as their structure and availability were consistent across clinical contexts. In contrast, flowsheet and laboratory features were modeled separately for ED and inpatient encounters due to differences in flowsheet types, hospital length of stay, ordering patterns, and availability between both settings. We extracted medical history from our EHR as a series of ICD-10-CM codes. We used published mapping algorithms to derive the Charlson comorbidity index (CCI) as well as indexed CCI components.^24^ We included all medications that were recorded in the EHR during the 90 days prior to stroke alert activation, which we categorized into therapeutic subgroups according to the Anatomical Therapeutic Chemical (ATC) classification, 2^nd^ level (2). To balance clinical granularity and high-dimensionality, we chose to represent medications by ATC2 and not ATC3 (which represents pharmacologic subgroups).

The sociodemographic variables we extracted included age, sex, race, ethnicity, and insurance type. We calculated age using the patient’s date of birth at the time of the stroke alert activation. Operational data included arrival method, type of person accompanying patient, and setting of the stroke alert. Physiologic flowsheet data were restricted to variables consistently documented in at least 50% of encounters in either ED or inpatient setting. The final set of retained flowsheets included heart rate, blood pressure, respiratory rate, oxygen saturation, temperature, emergency severity index (ESI), and pain assessments.

We chronologically aligned flowsheet data with stroke alert timestamps, excluding implausible values using predefined clinical thresholds. Given the irregular and episodic sampling nature of flowsheet data, we derived features capturing both chronic patterns and acute dynamics. Features included the mean and standard deviation over the full pre-alert window, rolling statistics over the final hour prior to alert, exponentially weighted moving averages (EWMA), and explicitly time-weighted summaries using a decay factor of 0.3.

Laboratory data from the 90 days preceding the alert were similarly stratified by clinical setting, and standardized and retained if present in at least 50% of encounters within either the ED or inpatient settings. Values were categorized as low, normal, or high using age-, and sex-specific institutional reference ranges. For each test, we extracted the most recent value to the stroke alert, as well as the median over the prior 30 and 90 days, the deviation between the last value from each median, the slope of the three most recent values, time since the last result, and counts of abnormal and normal results. When labs were repeated during the stroke alert encounter itself, we also calculated their median and standard deviation.

### Data Pre-processing

Following data extraction and cleaning, we pre-processed structured features to address missing values, standardize numeric variables, and encode categorical variables. We imputed continuous variables using an iterative modeling approach with multiple imputations (averaged across ten iterations). We imputed binary features using k-nearest neighbors (k = 5). We assigned an “Other” category for categorical variables to preserve informativeness. Continuous variables were standardized to zero-mean and unit variance, and categorical features were one-hot encoded.

### Feature Selection

To improve model performance and interpretability, we applied a two-step feature selection pipeline. We first used correlation-based filtering to remove highly collinear variables (i.e., those with |ρ| > 0.90), retaining the features with stronger outcome association. We then conducted recursive feature elimination with cross-validation (RFECV) using a gradient-boosted tree classifier (XGBoost), optimizing AUROC as the performance metric. An explicit AUROC threshold for feature exclusion was not set; instead, the RFECV process iteratively evaluated feature subsets and automatically selected the subset achieving the highest cross-validated AUROC, removing one feature at a time.

### AutoML Pipeline

We developed structured models using a customized automatic machine learning (AutoML) pipeline that systematically evaluated multiple algorithms, including Generalized Linear Models (GLMs), distributed random forests, gradient-boosted machines, extreme-gradient boosting (XGBoost) models, support vector machines, and ensemble models. The pipeline performed hyperparameter tuning for each model using grid search with five-fold cross-validation, and selected the model achieving the highest AUROC across folds for downstream analysis.

### Interpretability

To enhance interpretability and understand how our models generated predictions to ensure clinical relevance, we computed Shapley Additive Explanations (SHAP) to model predictions based on the held-out test set. SHAP analyses quantify the contribution of each feature to a model’s prediction, indicating whether it increases or decreases the likelihood of the outcome of interest. We visualized these effects using SHAP summary plots. For ensemble models, SHAP values were averaged across base learners to produce stable feature importance rankings.

### Multi-Modal Model Development

Each of the six component models was independently trained to represent a specific modality of clinical data: (SM1) a baseline model incorporating sociodemographic features, medication history, and Charlson Comorbidity Index (CCI); (SM2A) inpatient flowsheet data; (SM2B) ED flowsheet data; (SM3A) inpatient laboratory results; (SM3B) ED laboratory results; and (4) unstructured clinical notes (UsM). The decision to stratify flowsheets and laboratory data by care setting (ED vs. inpatient) reflects differences in clinical workflows and data availability across contexts.

We then constructed a multi-modal ensemble classifier that aggregated the logit outputs from each component model. We used a weighted averaging approach, where weights were proportional to each model’s F1 score. This allowed higher-performing models to contribute more heavily to the final prediction while retaining signal from all six modalities.

### Statistical Analysis; Model Evaluation

We stratified the stroke alerts by the presence or absence of ACD and compared the two groups using descriptive statistics of central tendency (mean or median) and variability (standard deviation [SD] or interquartile range [IQR]). We compared both groups using Student t-tests or Wilcoxon rank-sum tests for continuous variables, and chi-square or Fisher’s exact tests for categorical variables, as appropriate.

Model development was performed in Python 3.12.2, and all computations were conducted on NVIDIA V100 GPUs via the Mount Sinai Minerva HPC system.^25^ Model performance was evaluated using AUROC, sensitivity (Sn), positive predictive value (PPV), accuracy, specificity (Sp), and F1-score, all compared against the binary outcome. We did not determine area under the precision-recall curve (AUPRC) due to the overall balanced nature of the ACD labels. For LLM-based models, we trained all architectures for 100 epochs using an Adam optimizer. We selected the optimal MxBAI pooling strategy (mean, max, or combined) based on the highest AUROC, and identified the best-performing author type/timing combination from among the 12 variants (Supplementary Table 1). Structured models were trained using five-fold stratified cross-validation, and the model with the highest AUROC in each modality was selected for downstream integration.

## Results

Over the study period, we identified 16,512 stroke alerts, corresponding to 15,979 hospital encounters among 14,233 unique patients, reflecting that some patients had multiple stroke alert activations either during a single hospitalization or across multiple visits. Of these alerts, non-ACD was identified in 7,499 (45.4%) conditions and ACD was identified in 9,013 (55.6%). Most stroke alerts were activated in the ED (N=11,920; 72.2%). Among patients who had at least one stroke alert, the median age was 68.7 (IQR 57.0 - 79.7) years, and 9,017 (54.6%) were female. Compared with those whose alerts were not associated with ACD, patients with ACD-associated alerts were older (70.5 vs. 66.2 years, p<0.001), more likely to be male (47.9% vs. 42.4%, p<0.001), more frequently transferred from another acute care facility (4.1% vs. 2.5%, p<0.001), more often insured by Medicare (40.1% vs. 36.0%, p<0.001), and had a longer median hospital length of stay (3.8 vs. 1.3 days, p<0.001) (Table 1). Among patients with ACD, acute ischemic stroke was the most common subtype (59.0%), followed by transient ischemic attack (13.4%), intracerebral hemorrhage (7.3%), and other stroke types (Table 2).

**Table 1.**
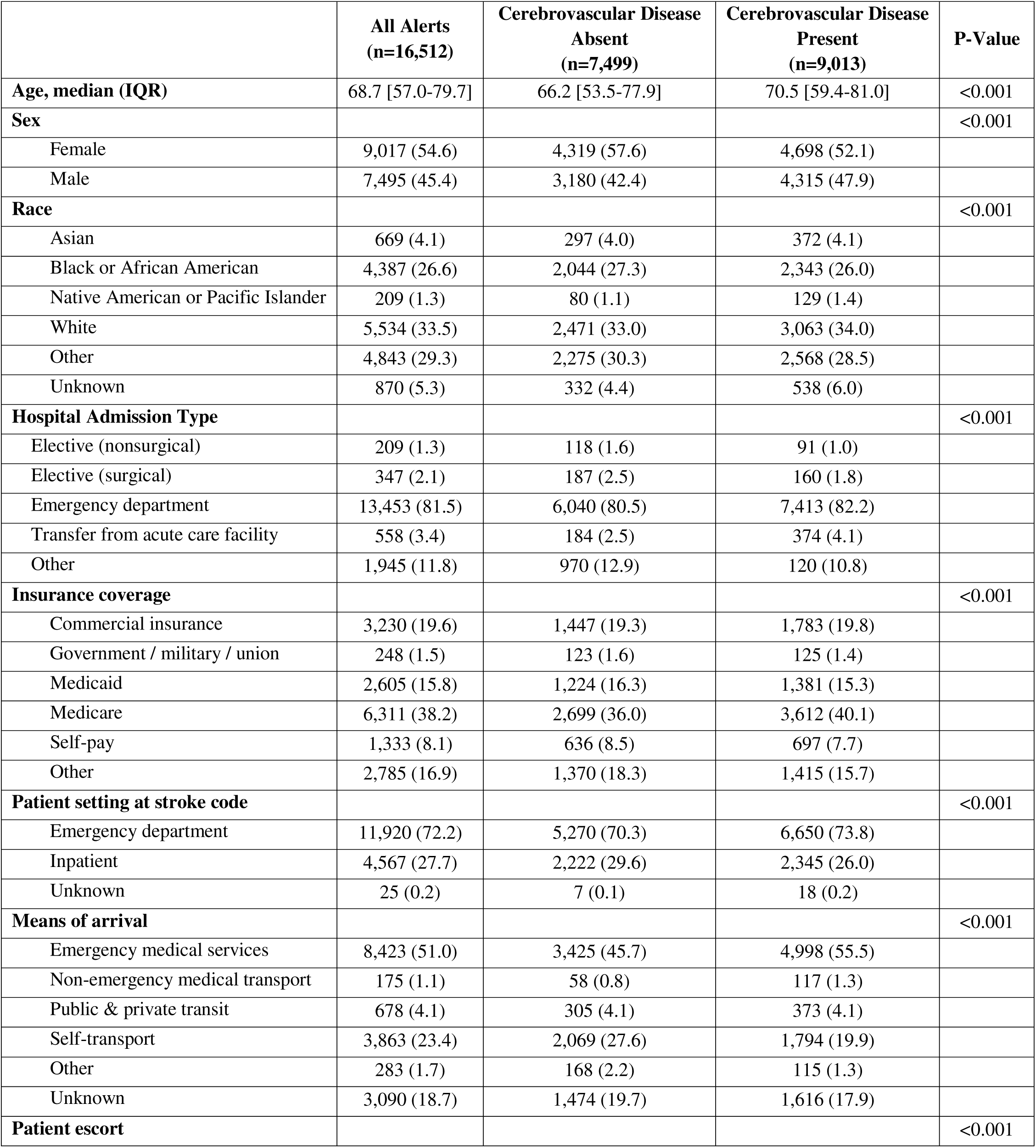
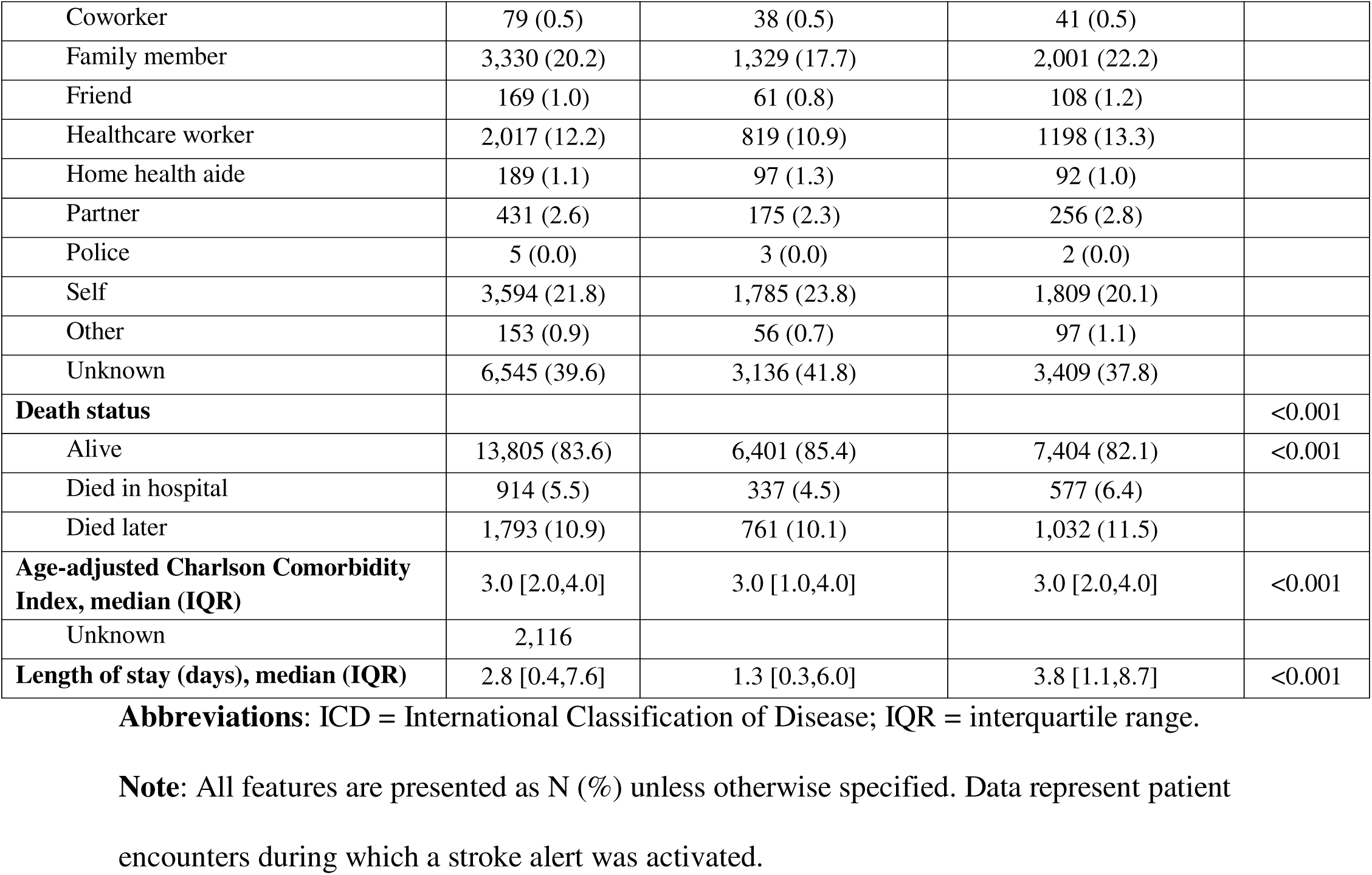
Baseline Characteristics of Patients by Cerebrovascular Disease Status.

**Table 2.** Distribution of Stroke Types Among Patients with Confirmed Cerebrovascular Disease.

| Stroke type | Cerebrovascular Disease<br>Present (n=9,013) |
| --- | --- |
| Acute ischemic stroke | 5,321 (59.0) |
| Transient ischemic attack | 1,210 (13.4) |
| Intracerebral hemorrhage | 659 (7.3) |
| Subarachnoid hemorrhage | 128 (1.4) |
| Subdural hemorrhage | 8 (0.0) |
| Other ICD codes | 1,687 (18.7)* |

**Abbreviations:** ICD = International Classification of Disease
**Note:** All values are presented as N (%).
\* “Other ICD codes” includes non-validated ICD codes or patients with cerebrovascular disease identified via radiology reports but without corresponding ICD codes.

### Unstructured Model (UsM): Clinical Notes

We tested 12 different configurations of note author types and timing to identify the optimal combination. The best-performing configuration was trial 10 (Supplementary Table 1). Among the 10,379 alerts with analyzable EHR notes, note characteristics are summarized in Supplementary Table 2.

The CLF model achieved an AUROC of 0.60, F1-score 0.62, PPV 0.57, and Sn 0.66. The BioWordVec model with mean pooling followed by logistic regression demonstrated an AUROC of 0.62, F1-score of 0.58, PPV 0.60, and Sn 0.56. The MxBAI model with mean pooling and FNN generated the highest overall performance among text-based models with AUROC 0.67, F1-score of 0.62, PPV of 0.65, and Sn 0.58. Consequently, this model was selected as the optimal text-based representation for downstream multimodal integration (Table 3). Performance metrics for this unstructured model, along with all structured models and the final ensemble model, are summarized in Table 4.

**Table 3.** Performance of Text-Based Classification Models.

| Model | Note set | Text handling | Accuracy | PPV | Sn | F1 | AUROC |
| --- | --- | --- | --- | --- | --- | --- | --- |
| Clinical-LongFormer | (10) | (built-in) | 0.58 | 0.57 | 0.66 | 0.62 | 0.60 |
| Feed-forward neural network | (10) | mxbai with mean pooling | 0.63 | 0.65 | 0.58 | 0.62 | 0.67 |
| Logistic Regression | (10) | BioWordVec with mean pooling | 0.59 | 0.60 | 0.56 | 0.58 | 0.62 |

**Abbreviations:** AUROC = Area Under the Receiver Operating Curve; F1 = Harmonic Mean of Precision and Recall; PPV = Positive Predictive Value; Sn = Sensitivity.
**Note:** Performance of models using unstructured clinical notes with different embedding strategies. Results shown for Clinical-LongFormer (CLF), a feed-forward neural network (FNN) using MxBAI embeddings, and logistic regression with BioWordVec embeddings. All models evaluated on the same test set.

**Table 4.** Performance of Models for Predicting Cerebrovascular Disease.

| Model | # Features | Data types | Accuracy | PPV | Sn | Sp | F1 | AUROC |
| --- | --- | --- | --- | --- | --- | --- | --- | --- |
| UsM* | NA | Clinical text notes | 0.63 | 0.65 | 0.58 | 0.63 | 0.62 | 0.67 |
| SM1† | 132 | Demographic information,<br>CCI, and medications | 0.61 | 0.59 | 0.95 | 0.20 | 0.73 | 0.69 |
| SM2-A‡ | 19 | IP flowsheets | 0.61 | 0.57 | 0.55 | 0.66 | 0.56 | 0.63 |
| SM2-B‡ | 18 | ED flowsheets | 0.60 | 0.62 | 0.66 | 0.52 | 0.64 | 0.62 |
| SM3-A‡ | 83 | IP labs | 0.62 | 0.58 | 0.62 | 0.62 | 0.60 | 0.65 |
| SM3-B§ | 24 | ED labs | 0.60 | 0.63 | 0.63 | 0.56 | 0.63 | 0.62 |
| MMM | 6 | Individual model logits | 0.67 | 0.68 | 0.76 | 0.56 | 0.72 | 0.72 |

**Abbreviations:** AUROC = area under the receiver operating curve; CCI = Charlson comorbidity index; ED = emergency department; IP = inpatient; MMM = multi-modal model; PPV, positive predictive value;; Sn, sensitivity; SM, structured model; Sp, specificity; UsM, unstructured model.
**Note:** Classification performance of six component models and one multimodal ensemble model. UsM represents the unstructured clinical notes model; SM1–SM3 refer to structured models trained on sociodemographics/medications/CCI (SM1), inpatient flowsheets (SM2A), ED flowsheets (SM2B), inpatient labs (SM3A), and ED labs (SM3B). The final row (MMM) represents the multimodal ensemble using logit-weighted integration. Best-performing classifier type is noted for each model.
\*Best model is MxBAI text handling with feed-forward neural network (FNN) classifier.
† Best model is an ensemble model.
‡ Best model is XGBoost.
§ Best model is logistic regression.

### Structured Data – Model 1 (SM1): Sociodemographics, Comorbidities, and Medications

Following feature selection using correlation and RFE, 132 features were retained for training. The best-performing SM1 was a stacking ensemble consisting of four base-learners (gradient boosting machines, XGBoost, GLM, and distributed random forest models), with GLM as the meta-learner, achieving an AUROC of 0.69, accuracy of 0.61, PPV of 0.59, recall of 0.95, specificity of 0.20, and an F1-score of 0.73.

SHAP analysis (Figure 3) demonstrated that older age, higher age-adjusted Charlson Comorbidity Index (CCI), EMS transport, a history of cerebrovascular disease, and transfer from an acute care facility were key features contributing to predictions toward the positive class (ACD). In contrast, self-transport and the use of certain medication classes—such as systemic antibiotics, antihistamines, antidepressants, and psychostimulants—tended to contribute more strongly toward the negative class (stroke mimic). Other variables, including insurance type, had less consistent influence on the model’s output.

**Figure 3.**
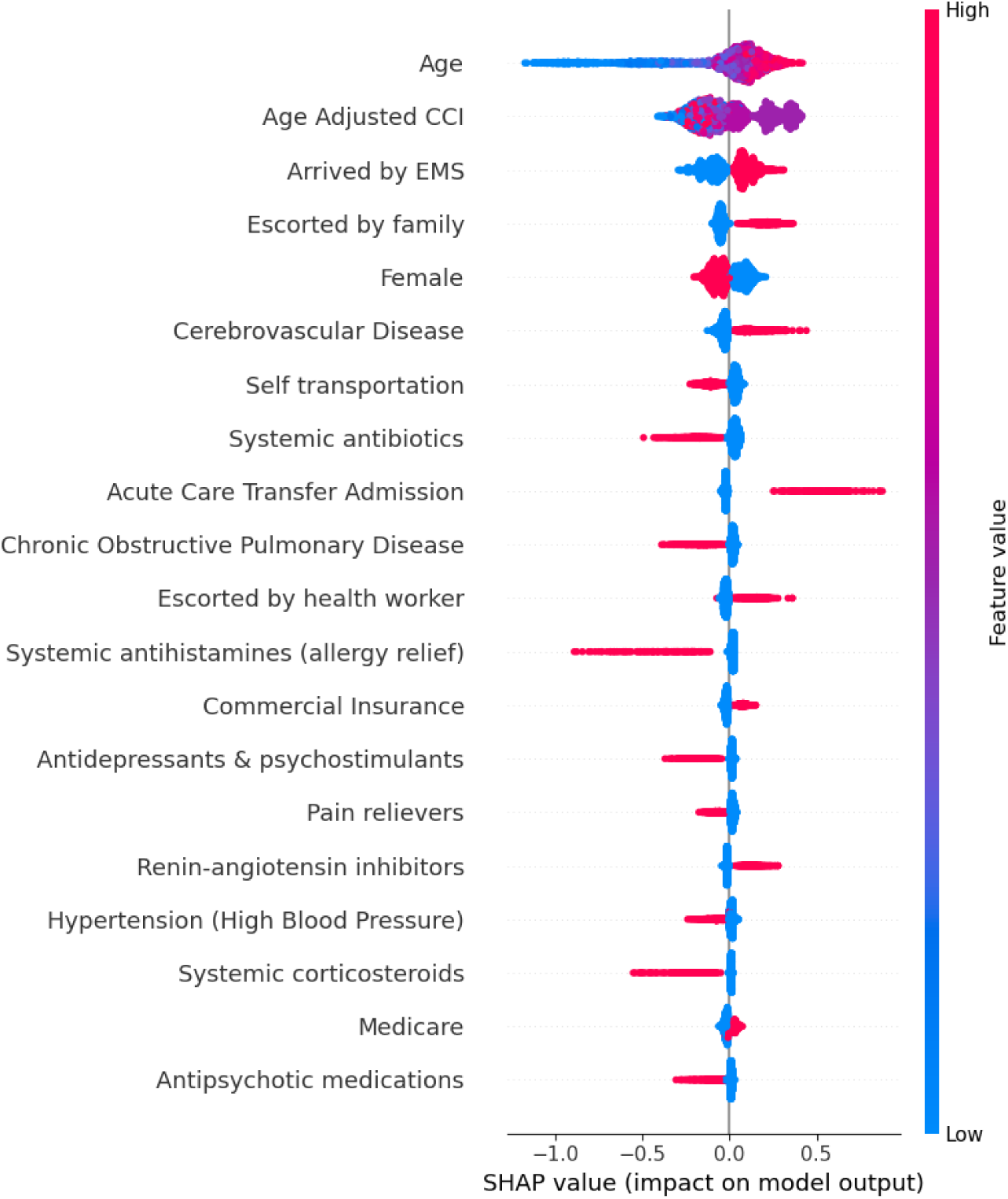
SHAP analysis for model SM1 (sociodemographics/medications/CCI) This plot displays the most influential features in SM1 (sociodemographic, medication and Charlson-Comorbidty Index [CCI]) contributing to model predictions of acute cerebrovascular disease. SHAP (Shapley Additive Explanations) values quantify the impact of each feature on the model’s output, indicating whether a given value increases or decreases the predicted likelihood of true stroke. Each dot represents a patient; color reflects the feature value (red = high, blue = low), and position along the x-axis reflects the direction and strength of that feature’s effect. This visualization allows for interpretation of global feature importance as well as individual prediction behavior.

### Model 2 (2A): Inpatient Flowsheets

Model 2A retained 19 features after feature selection and achieved an AUROC 0.63, accuracy 0.61, PPV 0.57, Sn 0.55, Sp 0.66, and an F1-score 0.56. SHAP analysis (Figure 4) revealed that higher diastolic and systolic blood pressures, elevated temperature, and increased respiratory rates were the most contributory to the positive class. Conversely, greater variability in pain scores, oxygen saturation, and pulse rate tended to shift predictions toward the negative class.

**Figure 4.**
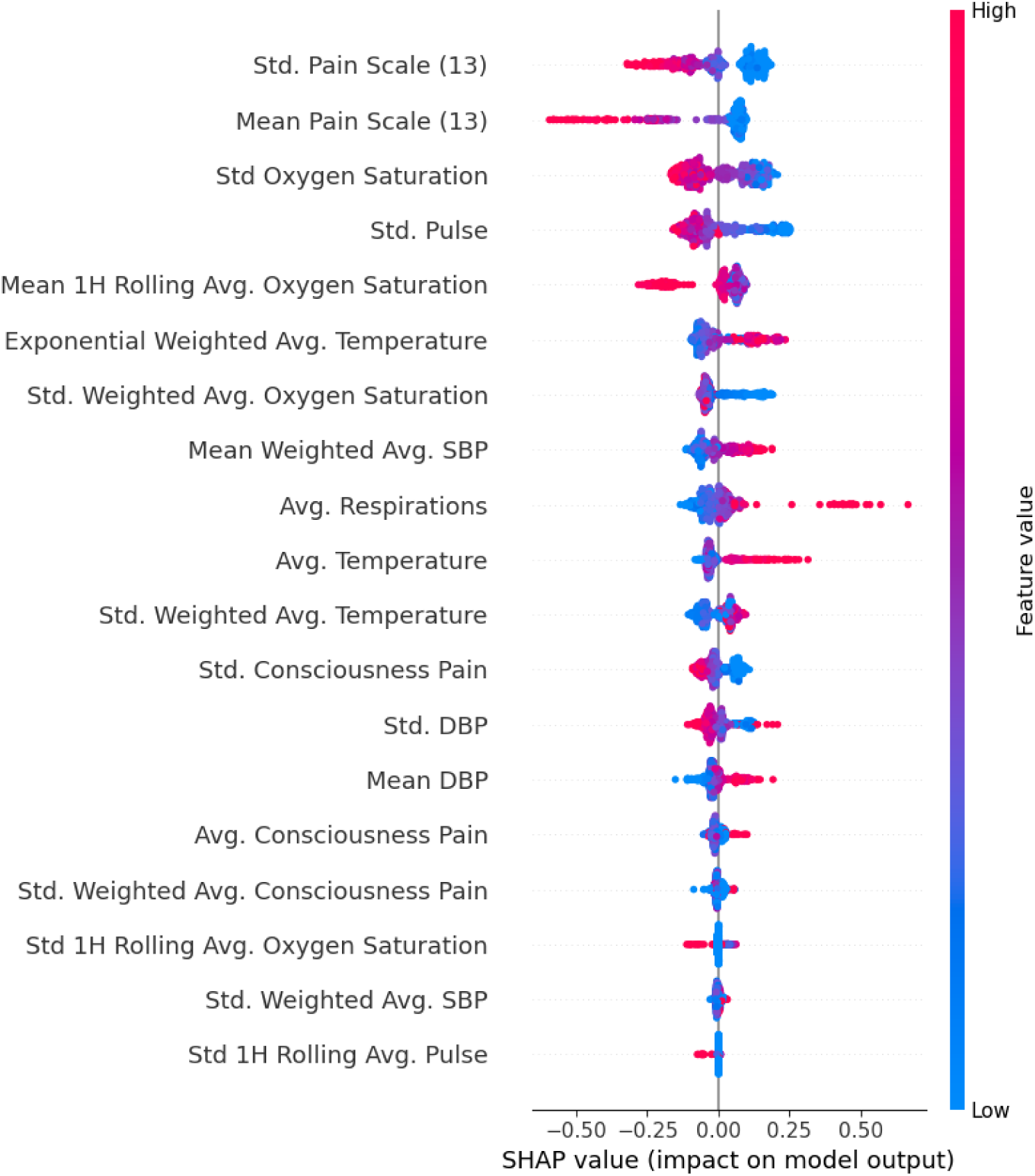
SHAP analysis for model SM2A (inpatient flowsheets) This plot displays the most influential inpatient vital sign and assessment features contributing to model predictions of acute cerebrovascular disease. SHAP (Shapley Additive Explanations) values indicate the direction and magnitude of each feature’s impact on the model’s output. Each dot represents a patient; color reflects the feature value (red = high, blue = low), and position along the x-axis indicates whether the feature increased or decreased the predicted probability of stroke.

### Model 2 (2B): ED Flowsheets

After feature selection, 18 features were retained in model 2B. XGBoost was the best-performing model, achieving an AUROC 0.62, accuracy 0.60, PPV 0.62, Sn 0.66, Sp 0.52, and an F1-score 0.64. SHAP analysis (Figure 5) showed that similar to Model 2 (A), higher systolic, diastolic, and greater variability in blood pressure were associated with predictions favoring the positive class. Additionally, Emergency Severity Index (ESI) was a strong predictor in the ED model, where better (i.e., higher) ESI scores were associated with stroke mimics. In contrast to Model 2(A), higher body temperature in the ED setting pushed predictions toward stroke mimic.

**Figure 5.**
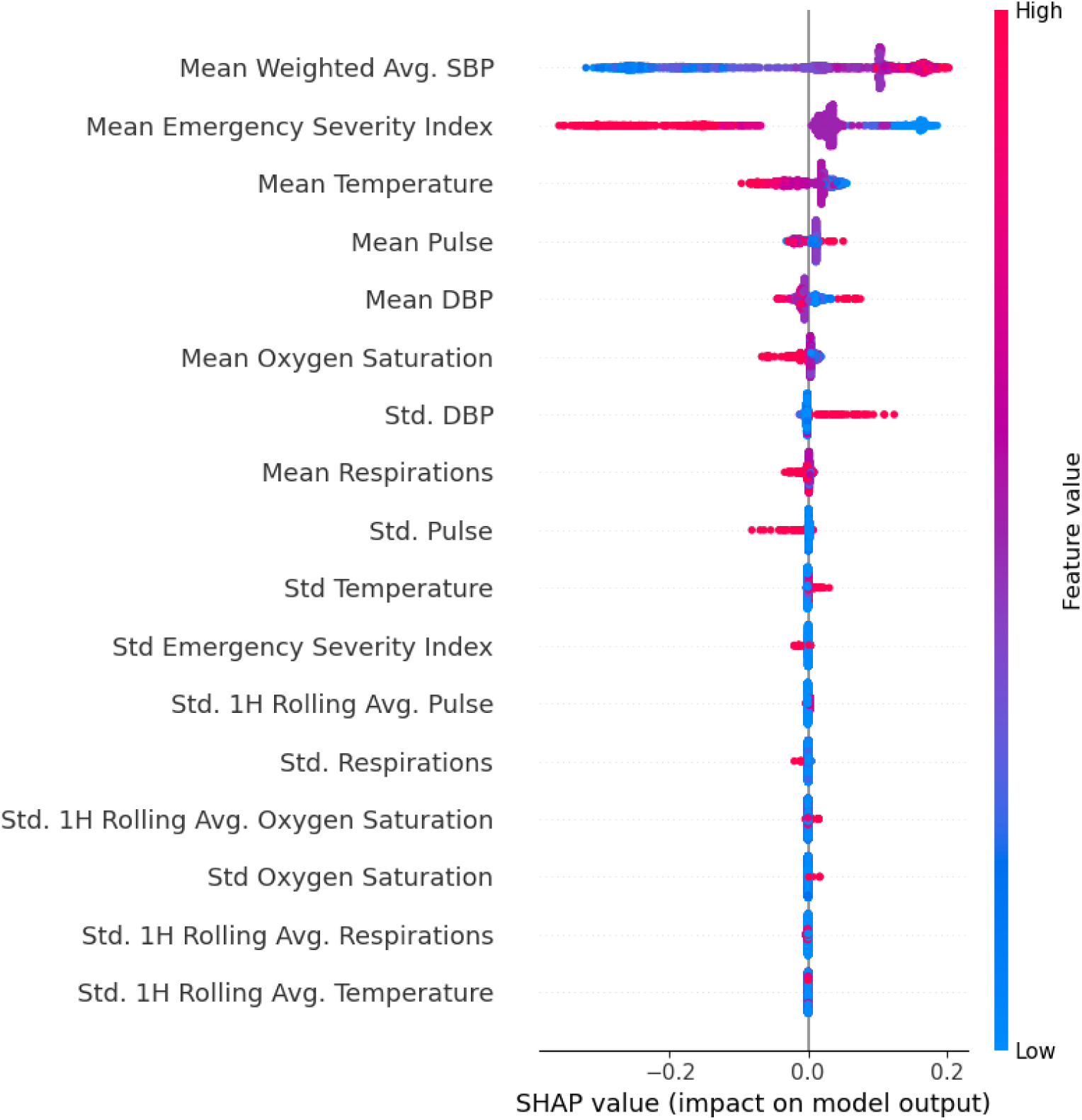
SHAP analysis for model SM2B (emergency department flowsheets) This plot displays the most influential emergency department vital sign and assessment features contributing to model predictions of acute cerebrovascular disease. SHAP (Shapley Additive Explanations) values indicate the direction and magnitude of each feature’s impact on the model’s output. Each dot represents a patient; color reflects the feature value (red = high, blue = low), and position along the x-axis indicates whether the feature increased or decreased the predicted probability of stroke.

### Model 3 (3A): Inpatient Laboratory Results

After feature selection model 3A retained 83 features, achieving the highest performance using XGBoost (AUROC 0.65, accuracy 0.62, PPV 0.58, recall 0.62, specificity 0.62, F1-score 0.60). SHAP analysis (Figure 6) revealed that deviations in red blood cell counts, differential lymphocyte percentage, elevated potassium over the preceding 90 days, and elevated carbon dioxide and creatinine levels during the stroke code encounter consistently contributed towards predictions for the negative class.

**Figure 6.**
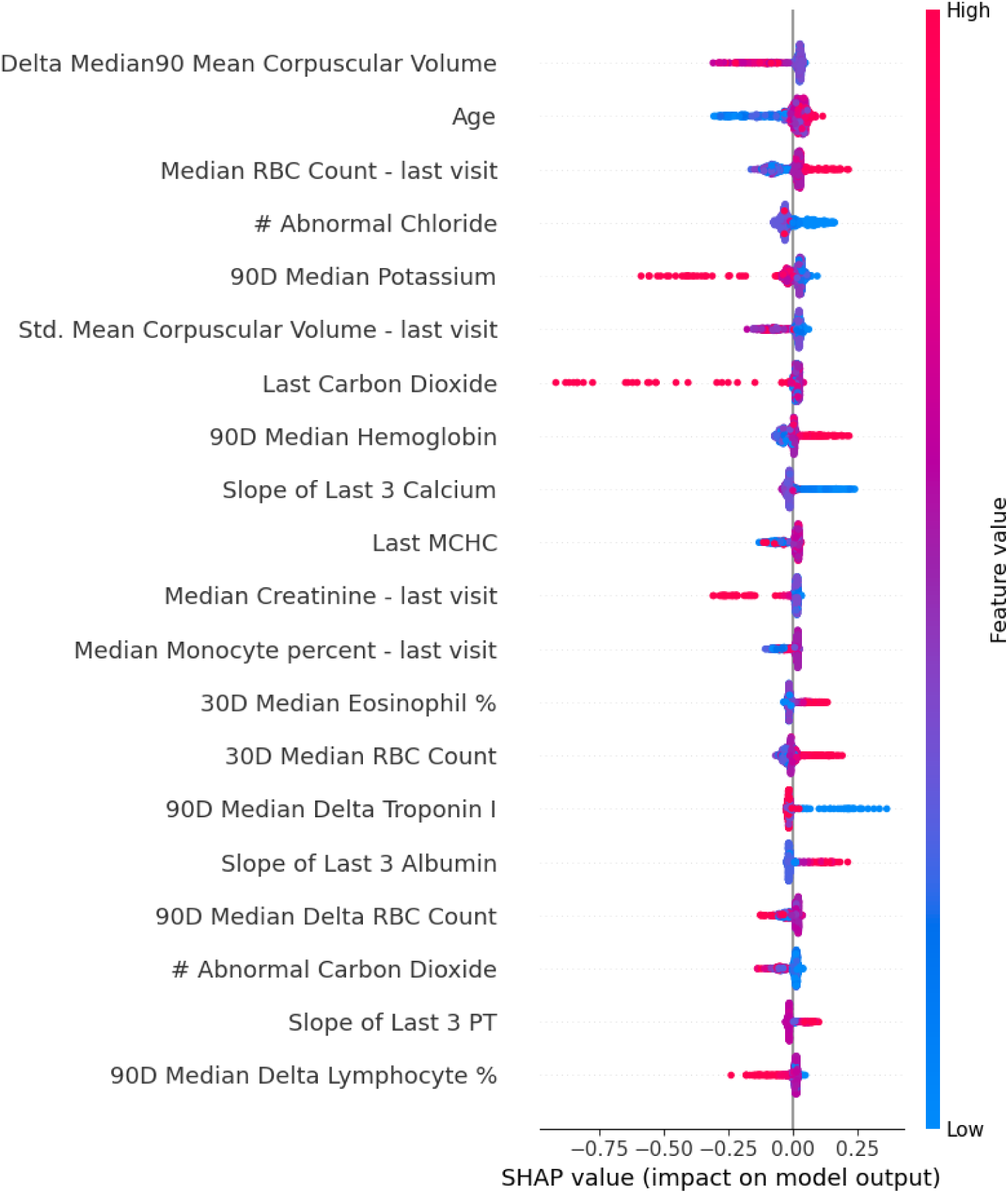
SHAP analysis for model SM3A (inpatient labs) This plot displays the most influential inpatient laboratory features contributing to model predictions of acute cerebrovascular disease. SHAP (Shapley Additive Explanations) values indicate the direction and magnitude of each feature’s impact on the model’s output. Each dot represents a patient; color reflects the feature value (red = high, blue = low), and x-axis position shows whether the feature increased or decreased the predicted probability of stroke.

### Model 3 (3B): ED Laboratory Results

Following feature selection, 24 features were retained in model 3B. Logistic regression was the best-perfrming model (AUROC 0.62, accuracy 0.60, PPV 0.63, recall 0.63, specificity 0.57, F1-score 0.63). SHAP analysis (Figure 7) showed that higher point-of-care glucose (either during the stroke encounter or over the last 30 days), higher absolute neutrophil count during the stroke encounter, and higher variation between mean platelet volume over preceding 30 days and that of the stroke encounter contributed strongly towards positive stroke prediction. By contrast, the total number of abnormal mean cell hemoglobin concentration (MCHC) test results in the preceding 90 days contributed towards a negative class prediction.

**Figure 7.**
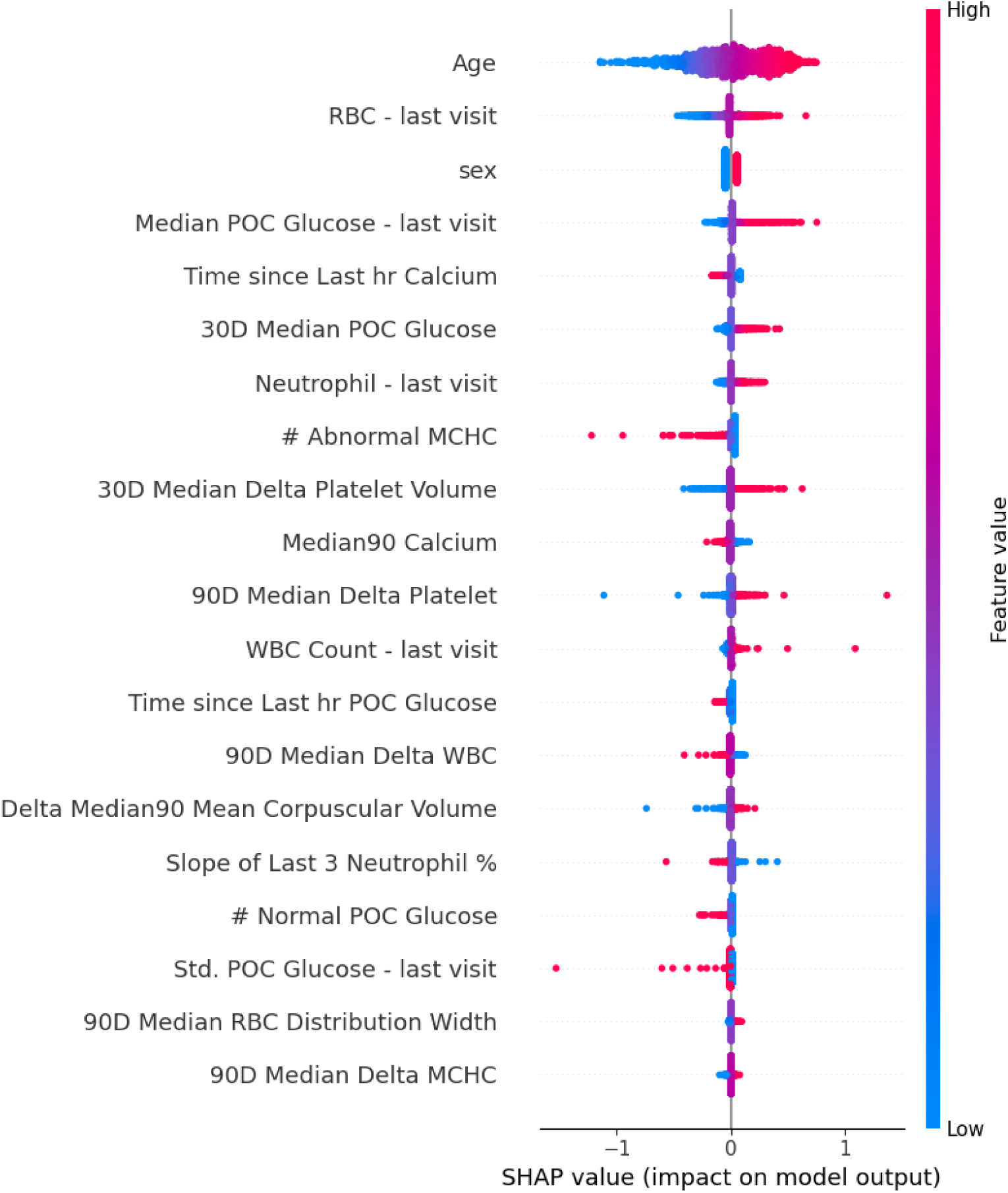
SHAP analysis for model 3B (emeregency department labs) This plot displays the most influential laboratory features from emergency department encounters contributing to model predictions of acute cerebrovascular disease. SHAP (Shapley Additive Explanations) values quantify each feature’s impact on the model’s output. Each dot represents a patient; color indicates the feature value (red = high, blue = low), and x-axis position reflects whether the feature increased or decreased the predicted probability of stroke.

### Final Model: (MMM) Multi-modal Model

The multimodal ensemble integrated outputs from all component models using weights proportional to their F1-scores. This final ensemble achieved an AUROC of 0.72, accuracy of 0.67, F1-score of 0.76, sensitivity of 0.72, and specificity of 0.56. The ensemble provided the most balanced performance, effectively combining structured and unstructured data insights to robustly predict cerebrovascular disease.

## Discussion

In this retrospective study of over 16,000 stroke alerts and 14,000 individual patients from a large, academic health network across 10 years, we applied ML and multi-modal modeling approaches to predict the presence of ACD at the time of stroke alert activation. More specifically, we developed predictive text models using LLM-derived (MxBAI, CLF) and static (BioWordVec) word embeddings from unstructured clinical notes, and applied an auto-ML approach to train a series of structured EHR data models based on (1) static structured data, and (2) labs and vital sign time series. We additionally combined structured and unstructured model outputs into a multi-modal ensemble classifier, which synergistically yielded the highest overall performance by AUROC.

We found that structured and LLM-based model performances were comparable. Furthermore, our multi-modal model demonstrated only moderate to good overall performance (0.72) and average specificity (0.56), suggesting potential avenues for improvement in our approaches to improve performance and reduce false-positive predictions. Despite this, our multi-modal model had a moderately high sensitivity (0.76), suggesting that this model would be less likely to identify patients with true cerebrovascular disease as mimics than vice-versa.

Several factors likely explain the overall performance ranges we found in our study. The most likely causative explanation is that the data types we used to train models UsM, SM2A/B, and SM3A/B (e.g., note text, laboratory values, and vital sign flowsheet data) likely did not possess highly distinctive patterns that permitted differentiation of both classes of patients. This is particularly likely for our textual models, which were based on clinical documentation written as as close as possible to the stroke alert, and included notes written by clinical staff across multiple disciplines – many if not most of whom lacked expertise in stroke (e.g., ED nurses and providers). However, the lack of distinctive patterns were also not specific to text data, given that we found that models SM2 and SM3 had roughly equivalent AUROC to model UsM’s in their ability to detect cerebrovascular disease at stroke activation.

Supporting this explanation is the notably high sensitivity we found for model SM1, which was trained on sociodemographic factors, medical comorbidities, and medications only. SM1’s superior discriminatory performance suggests that its training features contained more distinctive signal than individual notes, vital signs, or laboratory values to identify patients with true ACD as such (i.e., true positives). Despite its high sensitivity and resulting low false negative rate (5%), SM1 identified many false positives (i.e., stroke mimics identified as a stroke; 80%) and few true negatives (i.e., stroke mimics correctly identified as such; 20%).

In general, high-quality confirmatory tests often follow positive screening tests and are highly specific, whereas high-quality screening tests generally demonstrate high sensitivity—often at the cost of low specificity.^26,27^ In this fashion, SM1 can potentially be conceptualized as a “screen” for ACD. It is important to note that stroke alerts are operationalized exactly in this manner in many hospital systems, significantly contributing to excessive false-positive stroke alerts. Although in our study SM1’s performance was not adequate to solve this wide-spread problem, our results for SM1’s performance raise the interesting possibility of operationalizing this model as a potential automated replacement for human stroke alert activators. This would certainly require significantly further confirmatory studies.

The overall model performance metrics and AUROCs we found in our study are within reported ranges from prior studies that developed similar classifier models to distinguish acute stroke from stroke mimics.^12–20^ These studies range across a range of cohort types, settings, models, and input features. However, our overall model performances were notably lower than a recent analysis reported highly discriminant ML models from structured data and clinical note text to automatically identify acute ischemic stroke in EDs across a 13-hospital system in Pennsylvania.^19^ Despite the impressive performance reported in this work, our analysis possesses several notable differences that bear mention.

Because our analysis attempted to distinguish ACD among stroke mimics at the precise moment of stroke activation, our study design inherently incorporated a predictive timepoint which limited the inclusion of someinput features into our models. By contrast, this recent study incorporated all ED provider notes as predictive features at any point during their ED encounter.

By virtue of this, this prior approach may have included notes or other EHR data generated after a stroke diagnosis had been established and/or documented, thus increasing the model’s signal-to-noise ratio and potentially leading to excellent discriminatory performance. This difference has real-world implications. By limiting the inclusion of features by the stroke alert timestamp, our model generated a time-sensitive prediction that could potentially generalize to the real-world, commonly-encountered scenario of evaluating a patient with suspected (but unconfirmed) stroke. As such, our results establishes a foundation for an operationalizable model. While the AUROC achieved by our best-performing model was moderate, prospective validation via “silent pilot” (where front-line alert activators are unaware of the model predictions, but performance is nonetheless assessed in real-time. These approaches may further bolster our results, and potentially suggest an improved classifying ability than found in the present study.

Several limitations of our study warrant mention. First, our model training was conducted within a single health system, which may limit generalizability to other clinical settings. Second, we did not integrate raw imaging data or pre-hospital ambulance run sheet information, which may offer additional predictive value. However, imaging data is typically generated after the time of stroke code, which was felt to be too late for our desired prediction window. Third, we lacked data on the types of clinical staff (e.g., discipline, level of training, etc) activating the stroke alerts as well as the specific clinical reasons that resulted in stroke alert activation. Such data is very likely to distinguish both patient classes, but is not recorded as part of the stroke alert workflow at our institution.

Fourth, we defined “ACD” as a composite of multiple disease states (acute ischemic stroke, intracerebral hemorrhage, transient ischemic attack, subarachnoid hemorrhage). However, given that these states often present similarly, i.e., with focal neurological symptoms, we felt that it was unlikely for a disease-specific predictive model to perform better than a composite model. Fifth, we combined data from alerts originating both inpatient and ED settings, which have different data type and patient profiles. However, we did train models 2A/B and 3A/B using data from ED and inpatient settings separately, incorporating the output from these setting-specific models into our final multi-modal classifier. Sixth, as occurs in many retrospective studies of EHR data, many variables had missing data.

Given the differences in data types between ED and inpatient settings, future work will seek to develop multi-modal models that are specific to either ED and inpatient populations. Additionally, future studies will seek to prospectively validate our findings at our institution, as well as in external populations and documentation practices. Finally, additional avenues for exploration include analyzing neurological consultation notes to inform efforts at redesigning stroke alert activation pathways. Our findings suggest that the information currently documented by front-line staff (who are often not neurology-trained) may be insufficient to reliably and rapidly distinguish ACD. If implemented in a real-world clinical setting, the model would ideally run automatically on EHR data without user input, with outputs intended for interpretation by clinicians within existing stroke alert workflows.

## Conclusion

This study highlights the potential of ML in developing automated tools for accelerating distinction between stroke and non-stroke in the hyperacute setting. Ensemble approaches combining structured data and LLM-based models demonstrated superior classifying performance than either data type alone, underscoring the need for multimodal approaches to improve stroke alert specificity and optimize resource allocation in acute stroke care. Prospective studies are needed to validate our findings, but also develop text-based approaches for redesigning stroke protocol evaluations by non-neurology clinicians.

## Acknowledgments

None.

