## Supplementary material for "Predicting Acute Cerebrovascular Events in Stroke Alerts Using Large-Language Models and Structured Data": Supplental Materials

**Supplementary Table 1**. Text permutation experiments

| **Text Permutation** | **Note type criteria** |
| --- | --- |
| 1 | All notes |
| 2 | All notes – earliest notes only (first 30 min) |
| 3 | All notes – latest notes only (last 30 min) |
| 4 | All ED notes only |
| 5 | All ED notes – earliest only |
| 6 | All ED notes – latest only |
| 7 | ED notes - triage RN notes excluded |
| 8 | ED notes - triage RN notes only |
| 9 | ED notes, H&Ps, and progress notes |
| 10 | H&P, event, progress, attestation, transfer notes (ED and non-ED) |
| 11 | notes from set (10) – earliest only |
| 12 | notes from set (10) – latest only |

Abbreviations: ED, emergency department; ; H&P, history and physical; RN, registered nurse.

**Supplementary Table 2.** Characteristics of Notes Included in Textual Models

|  | **All stroke alerts**  **(N=10,379) ^*^** | **ED alerts only**  **(N=7,837)** | **Inpatient alerts only**  **(N=2,542)** |
| --- | --- | --- | --- |
| **Total discrete notes** | 159,260 | 14,774 | 144,133 |
| **Notes per alert** | 2 (1 - 4) | 1 (1 – 2) | 24 (9 – 56) |
| **Characters per note** | 1,536 (303 - 4,923) | 125 (76 – 205) | 1,966 (431 – 5319) |
| **Words per note** | 281 (58 – 904) | 44 (26 – 56) | 357 (81 – 981) |
| **Sentences per note^†^** | 45 (8 – 191) | 3 (1 – 5) | 64 (12 – 206) |
| **Time from note filing to stroke alert, minutes** | 9,607.0  (2,478.0 – 28,560.0) | 22.2  (7.2 – 79.2) | 11,640  (3,900 – 32,000) |
| **Notes per encounter, by author category^‡^** |  |  |  |
| Provider | 8.6 ± 40.1; 0 (0–2) | 0.30 ± 1.11; 0 (0–0) | 34.0 ± 75.5; 13 (5–28) |
| Nurse | 3.5 ± 8.2; 1 (0–2) | 1.6 ± 1.15; 1 (1–1) | 9.4 ± 15.0; 5 (2–9) |
| Other | 3.3 ± 19.7; 0 (0–0) | 0.02 ± 0.16; 0 (0–0) | 13.4 ± 38.1; 3 (0–13) |
| **Number of alerts with specific note types^§^** |  |  |  |
| ED Triage/Intake | 9,126 (87.9%) | 7,725 (98.6%) | 1,388 (54.6%) |
| ED Notes | 2,941 (28.3%) | 1,623 (20.7%) | 1,313 (51.7%) |
| Progress Notes | 2,271 (21.9%) | 72 (0.9%) | 2,194 (86.3%) |
| ED Provider Notes | 2,120 (20.4%) | 808 (10.3%) | 1,307 (51.4%) |
| History & physical | 1,809 (17.4%) | 38 (0.5%) | 1,768 (69.6%) |
| Consults | 1,623 (15.6%) | 34 (0.4%) | 1,585 (62.4%) |
| Event Note | 1,621 (15.6%) | 30 (0.4%) | 1,588 (62.5%) |
| ED Progress Notes | 1,536 (14.8%) | 490 (6.3%) | 1,044 (41.1%) |
| Attestation | 1,017 (9.8%) | 77 (1.0%) | 937 (36.9%) |
| Transfer of Care | 713 (6.9%) | 1 (0.0%) | 710 (27.9%) |

**Abbreviations**: ED, emergency department.

**Note**: Continuous variables (e.g., notes per alert, characters per note, words per note, sentences per note, time from filing to alert) are reported as medians (IQR).

^*^ Out of 16,512 stroke alerts identified in our registry by ScanNER framework, only 10,379 had analyzable EHR notes.

† Sentences were counted using the NLTK sentence tokenizer.

‡ For note authors, values represent the average (mean ± SD) and median (IQR) numbers of notes authored per encounter, stratified by provider, nurse, and other categories. Providers included physicians, residents, fellows, physician assistants, and nurse practitioners; nurses included registered nurses, licensed practical nurses, and nurse anesthetists; other included ancillary staff such as social workers, nutritionists, and therapists.

§ Counts for specific note types represent the number of stroke alerts with at least one note of that type. Because encounters often contained multiple note types, totals may exceed 100%.
